# Survival After In-Hospital Cardiac Arrest and Return of Spontaneous Circulation: An Exploration of Outcome Variation and Relationship to Hospital Area Social Deprivation

**DOI:** 10.64898/2025.12.17.25342524

**Authors:** Luke Andrea, Thomas Butler, Ari Moskowitz

**Affiliations:** Bronx Center for Critical Care Outcomes and Resuscitation Research, Montefiore Medical Center, the Bronx, NY; Division of Critical Care Medicine, Montefiore Medical Center, the Bronx, NY

**Author notes:** Corresponding Author: Luke Andrea, MD MS. **Luke Andrea, MD MS**, **Thomas Butler**, MS, **Ari Moskowitz, MD MPH**.

## Abstract

**Background:** In-hospital cardiac arrest (IHCA) survival has improved over the past two decades, resulting from better acute resuscitation survival. Post-resuscitation care is a key link in the IHCA chain-of-survival, yet post-resuscitation survival has remained stagnant over time.

**Hypothesis:** We hypothesized substantial hospital-to-hospital variation exists in risk-standardized post-resuscitation survival rate (RSSR) and hospital-area social deprivation is associated with worse RSSR.

**Methods:** We performed a cohort study of the American Heart Association Get With The Guidelines^®^-Resuscitation registry, linked to the American Hospital Association survey. We included adult IHCA patients with sustained return of spontaneous circulation (ROSC) from 2001–2024. Post-resuscitation RSSR was the ratio of the predicted (hospital-specific average of patient-level predictions from mixed-effects model) to expected (adjusted population-averaged predictions) survival, multiplied by the unadjusted full-cohort survival. Social deprivation index (SDI) was assigned by ZIP-to-ZCTA linkage and analyzed by quartile. Hospitals lacking ZCTA linkage were excluded from SDI analyses.

**Results:** Of 686,273 IHCA, 206,467 from 755 hospitals were included for primary analysis. Overall, 71,691 (34.7%) patients who achieved ROSC survived to discharge. Median RSSR was 33.9% (IQR 32.6-35.1%). Variation in RSSR across hospitals was substantial—ranging from 25.0 to 44.8%. A total of 595 (78.8%) hospitals were linked by zip-code to SDI. Patients at hospitals in the lowest quartile of SDI (least deprivation) had higher post-resuscitation RSSR compared to patients in the highest quartile (aOR 1.13, 95% CI 1.03-1.23, p<0.01), although no monotonic relationship existed between hospital SDI quartile and RSSR quartile. Hospitals in higher SDI quartiles (more deprivation) had higher proportions of early post-resuscitation fever and death.

**Conclusions:** Substantial hospital-to-hospital variation exists in post-resuscitation survival, and greater community social deprivation predicts worse post-resuscitation outcomes at the patient level. These results identify the post-resuscitation phase of care as a promising area for future quality improvement and research efforts to improve outcomes after IHCA.

**Summary:** With the Get With The Guidelines^®^-Resuscitation registry (2001-2024) we evaluated adults with sustained return of circulation after in-hospital cardiac arrest to quantify hospital-level variation in risk-standardized post-resuscitation survival and explore whether hospital-area social deprivation is associated with worse post-resuscitation outcomes.

**Author Statement:** This is original work from the authors, has not been published previously, and is not under consideration elsewhere.

**Approval:** This work was approved by the Albert Einstein College of Medicine Institutional Review Board (2025-16756) with exempt status.

**Conflicts of Interest:** There are no conflicts of interest for any authors.

**Funding Information:**

– Luke Andrea: Grant from the Clinical and Translational Science Award (CTSA) program, funded by the National Center for Advancing Translational Sciences (NCATS) at the National Institutes of Health (NIH) grant number K12TR004411 to perform research unrelated to this project.
– Ari Moskowitz: Grant from the National Institutes of Health/National Heart, Lung, and Blood Institute (4R33HL162980) to perform research unrelated to this project; Volunteer member of the ILCOR Advances Life Support Task Force; Volunteer Member of the American Heart Association Post-Cardiac Arrest Care Guideline Writing Group.
– Authors Luke Andrea and Ari Moskowitz are supported by an award from the American Heart Association (24GWTGDRA1308863).

## Introduction

In-hospital cardiac arrest (IHCA) is a major threat to cardiovascular health, impacting over 290,000 individuals in the United States each year.^1,2^ Overall survival outcomes for in-hospital cardiac arrest (IHCA) have improved over the past decade,^3–7^ largely due to improved rates of acute resuscitation survival (i.e. more return of spontaneous circulation (ROSC)). Among initial survivors of cardiac arrest, however, post-arrest morbidity and mortality remain high with many patients suffering long-term neurological, functional, and psychiatric disability.^4,6–8^ Efforts to improve IHCA outcomes have primarily focused on acute resuscitation, with less intensive efforts dedicated to understanding and improving post-resuscitation care.^9^ Highlighting this, the American Heart Association (AHA) Get With The Guidelines^®^ Resuscitation (GWTG^®^-R) Hospital Recognition Criteria only include intra-arrest benchmarks.^10,11^

Factors associated with social determinants of health may impact post-resuscitation survival and contribute to persistent disparities in IHCA outcomes.^12^ In a landmark study using the GWTG^®^-R registry, Joseph et. al. found that the survival gap between Black and White IHCA victims narrowed substantially over time, and that the improvement in outcomes for Black as compared to White IHCA victims was largely driven by improvements in acute resuscitation survival with little change in post-resuscitation survival.^13^ Guideline adherent post-resuscitation care, including temperature control, coronary intervention, extracorporeal support, and neuroprognostication, can be costly and require a high degree of expertise. Whether post-resuscitation survival is associated with hospital area social deprivation is unknown.

To date, hospital-to-hospital variation, and contributors to disparity in post-ROSC survival have not been comprehensively explored. Leveraging the large, nationwide, and prospectively collected GWTG^®^-R registry, this study adds new insights into survival and disparities after successful IHCA resuscitation.

## Materials and Methods

### Study Design and Data Source

This is an observational study of the prospectively collected GWTG^®^-R database provided by the American Heart Association (AHA), previously described in detail.^14^ Hospitals contributing to the database submit clinical information of consecutive patients with IHCA using an online case report form and Patient Management Tool™ (IQVIA, Parsippany, New Jersey). Data on participating hospitals was provided by the American Hospital Association Annual Survey.

Participating GWTG^®^-R hospitals are required to adhere to local regulatory and privacy guidelines. The Institutional Review Board (IRB) at the Albert Einstein College of Medicine deemed the use of the de-identified GWTG^®^-R as non-human subjects research (IRB#2025-16756).

### Study Population

This study included adult patients (≥18 years of age) who suffered IHCA and achieved sustained ROSC (defined as a spontaneous circulation for at least 20 minutes) during the years 2001 through 2024. We excluded non-index cardiac arrests, arrests that did not occur in an intensive care unit or hospital ward, and arrests where information on survival outcomes was missing. Patients over the age of 122 were excluded as data entry errors. Arrests at hospitals contributing fewer than 10 events to the database over the study period were excluded.

### Social Deprivation Index

Social Deprivation Index (SDI) is a composite of income, education, employment, housing, household characteristics, and transportation calculated by the Robert Graham Center for Policy Studies in Family Medicine and Primary Care.^15,16^ The score ranges from 0 to 100, and higher numbers represent greater social deprivation. For this data, the aggregated SDI American Community Survey was linked to the American Heart Association GWTG^®^-R registry. SDI data was associated to hospital data through Zip Code Tabulation Areas (ZCTA). As the GWTG®-R database provides Zip Codes and not ZCTAs, an established bridging dictionary was used (acs-aggregate/crosswalks/zip_to_zcta/zip_zcta_xref.csv at master · censusreporter/acs-aggregate · GitHub).^17^ This dictionary data associates ZCTA with Zip codes based on latitude and longitude, a method that has been employed previously for epidemiological research.^18,19^

### Outcome Measures

The primary outcome was survival to hospital discharge. Additional exploratory outcomes included fever recorded in the 24-hours after ROSC, and early death after successful resuscitation defined as death in ≤72-hours after ROSC. These latter, process outcomes were selected to reflect guideline-recommended care process using available post-resuscitation variables in the GWTG^®^-R database.^20,21^ Neurologically favorable survival was not included as an outcome measure given the high degree of missingness for IHCA survivors in the GWTG^®^-R registry.

### Statistical Analysis

#### General Cohort Characteristics

Demographic, arrest, and hospital characteristics are reported both overall and separated for post-resuscitation survivors and non-survivors. Categorical variables are presented as counts and percentages. Continuous variables are presented as means with standard deviations (SD) or medians with interquartile ranges (IQR) as appropriate. For all analyses the years 2001 and 2002 were combined owing to a low number of cardiac arrests in the registry in 2001.

#### Hospital Variation in Post-Resuscitation Outcomes

Risk-standardized post-resuscitation survival rate (RSSR) at each hospital was calculated as the ratio of the predicted to expected outcome, multiplied by the unadjusted outcome for the full cohort. Predicted outcome was defined as the hospital-specific average of patient-level predictions within each hospital system from a mixed-effects logistic regression model, while incorporating both fixed effects and the hospital system’s random intercept. Expected outcome was calculated from a logistic regression model, with the same covariates (excluding hospital system), using population-averaged predictions adjusted for the overall cohort’s covariate distribution. The regression models included the fixed effects of arrest year, patient age, sex, race, ethnicity, illness category, comorbidities and pre-arrest conditions [congestive heart failure, history of congestive heart failure, myocardial infarction, history of myocardial infarction, respiratory insufficiency, metabolic or electrolyte abnormality, baseline depressed neurologic status, stroke, CNS non-stroke, pneumonia, trauma, dialysis/extracorporeal filtration therapy ongoing, renal insufficiency, hepatic insufficiency, sepsis, malignancy, hypotension/hypoperfusion, diabetes], pre-arrest vasopressor use, pre-arrest mechanical ventilation] hospital characteristics [hospital size, hospital teaching status, geographic location, urban vs. rural location, hospital ownership], and arrest characteristics [initial rhythm, arrest location, witnessed status, day or night arrest timing, weekend or weekday arrest timing, and arrest duration]. In a post-hoc sensitivity analysis, the model was additionally adjusted for COVID-19 status.

For the primary analysis of hospital-to-hospital variation in risk-standardized outcomes, hospitals were ordered from lowest performing to highest performing on a caterpillar plot. To show changes in hospital performance over time, hospitals were placed in quartiles of performance within the time periods of 2001-2012 and 2013-2024, and an alluvial plot was then used to demonstrate changes in the quartile of hospital performance between the two time periods. Only hospitals present in the database during both time periods were included in this data visualization. For each quartile of RSSR, the quartile average ROSC rate is additionally provided to examine whether differential ROSC rates—with higher performing hospitals achieving ROSC in more critically ill patients and thus resulting in a low post-resuscitation survival—potentially contribute to variation in post-ROSC survival.

#### Patient-Level Predictors of Post-Resuscitation Survival

The mixed-effects regression model described above was used to determine patient-level predictors of post-resuscitation survival. For this analysis hospital was treated as a random intercept to account for clustering within hospitals.

#### Relationship between Social Deprivation Index and Post-Resuscitation Survival

Two approaches were used to assess the relationship between SDI and post-resuscitation outcomes. For these analyses, only hospitals in the database that could be linked to SDI were included. First, at the patient level, a mixed-effects logistic regression model was used with post-resuscitation survival as the binary outcome variable, fixed effects as above used in the risk-standardization model with hospital as a random effect, and including an additional fixed effect of hospital SDI quartile as the primary exposure of interest. Second, we plotted hospital RSSR across SDI quartiles using boxplots and assessed the monotonic association between SDI quartile (ordinal) and continuous RSSR with Spearman’s rank correlation.

#### Post-resuscitation Care Processes

As an exploratory analysis of potential contributors to variation in post-resuscitation survival, the occurrence of fever in the first 24 hours after ROSC and death within 72 hours of ROSC were assessed. As temperature after ROSC is an optional variable in the database, hospitals where >50% of patients were missing temperature data were excluded from the fever analysis. Patients missing ROSC date or discharge date were not included in the early death analysis. Hospital-averaged rates of fever and early death were separately plotted against quartiles of hospital RSSR and quartiles of hospital SDI and assessed for monotonic trend using Spearman’s rank correlation test. As death within 24 hours is a competing risk for fever within 24 hours, patients who died within 24 hours were categorized as having a fever. In a sensitivity analysis, patients who died within 24 hours were categorized as not having a fever.

Statistical analyses were performed and figures were generated using R statistical software on the American Hospital Association Precision Medicine Platform. Rates of data missingness for all included variables can be found in Supplement Table 1.

## Results

### Cohort Selection and Characteristics

A total of 686,273 patients were present in the full cohort, and 206,467 patients included in the primary analysis cohort (Figure 1). Characteristics of the primary analysis cohort are presented in Supplemental Table 2. Patients in the final cohort were drawn from 755 hospitals, with numbers of patients included from each hospital shown in Supplemental Figure 1.

**Figure 1:**
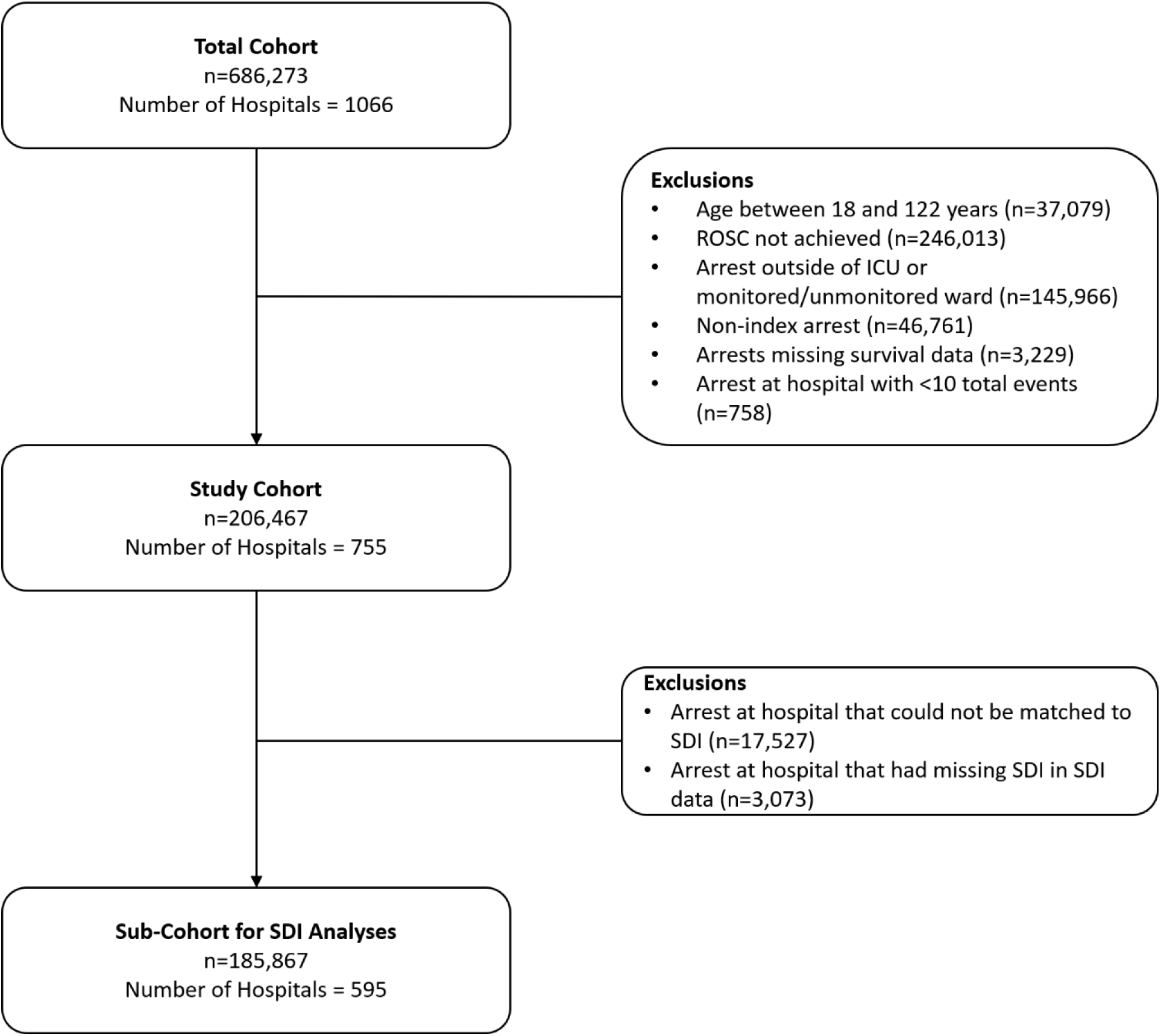
Study Flow Diagram.

### Hospital Variation in Risk-Standardized Post-resuscitation Survival Rate (RSSR)

A total of 71,691 (34.7%) patients survived to hospital discharge. RSSR ranged from 25.0-44.8%, with a median of 33.9% (IQR 32.6-35.1%). Substantial variation in RSSR is demonstrated in Figure 2A (and with COVID adjustment in Supplemental Figure 2). A Sankey plot illustrating hospital quartile classifications of RSSR across two time periods is shown in Figure 2B, demonstrating some hospitals with stability over time but many with changes in performance quartile across the two time periods.

**Figure 2:**
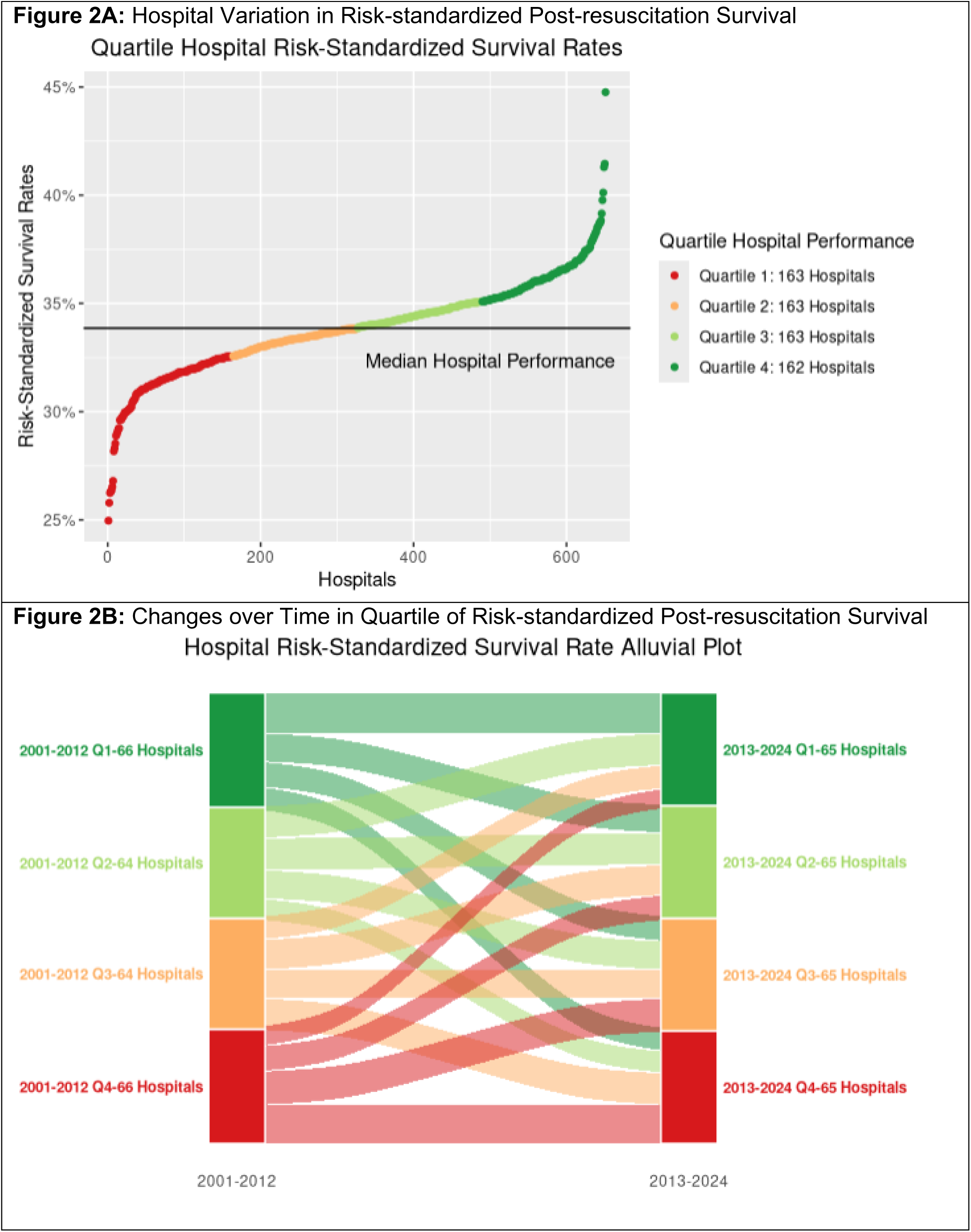
Hospital Risk-Standardized Post-Resuscitation Survival Characteristics.

The average rate of return of spontaneous circulation among hospitals in the highest quartile of RSSR was 60.6%. Remaining quartiles had average ROSC rates of 58.8%, 58.8%, and 60.2% (lowest quartile), respectively.

### Patient Level Predictors of Post-resuscitation Survival

Select predictors of post-resuscitation survival from the patient-level regression model can be found in Figure 3, the results for all covariates are in Supplemental Table 3. Older age, underlying malignancy, hepatic insufficiency, Black race, Hispanic ethnicity, longer arrest duration, pre-arrest receipt of vasopressors, and pre-arrest receipt of invasive mechanical ventilation were among the strongest predictors of worse post-resuscitation survival. Changes over time in post-resuscitation survival have been mixed with an initial period of relative stability between 2001 and 2009, followed by modestly lower rates from 2010 through 2019, and then a larger fall in post-resuscitation survival during the COVID-19 pandemic in 2020 and 2021. Since COVID-19, post-resuscitation survival has modestly improved but not yet back to the pre-pandemic rates.

**Figure 3:**
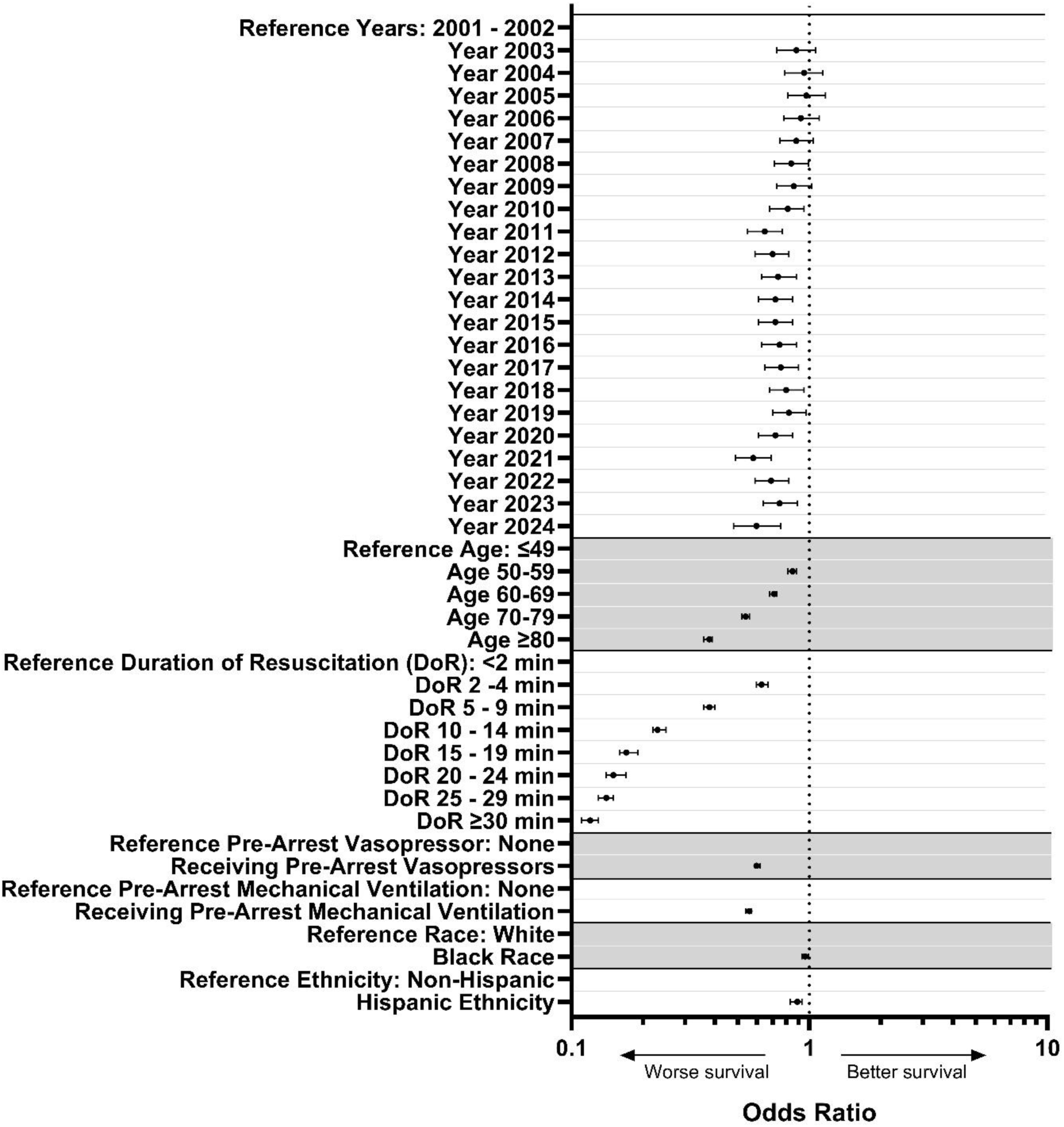
Forest Plot of Select Patient-Level Predictors of Post-Resuscitation Risk-standardized Survival.

### Social Deprivation Index and Risk-Standardized Post-resuscitation Survival Rate

A total of 595 (78.8%) hospitals in the cohort could be linked by zip code to SDI. Median SDI was 64.0 (IQR 40.5-83.0; higher scores indicating greater deprivation). Hospitals in the lowest quartile of SDI (least deprivation) had a median RSSR of 33.9% (IQR 32.5-35.2%), with subsequent quartiles having median RSSR of 33.7% (IQR 32.6-34.8), 34.0% (IQR 32.6-35.2%), and 33.8% (IQR 32.6-35.1%), respectively (Figure 4, and with COVID adjustment in Supplemental Figure 3). Higher quartiles of SDI were not associated with lower RSSR (p=0.79).

**Figure 4:**
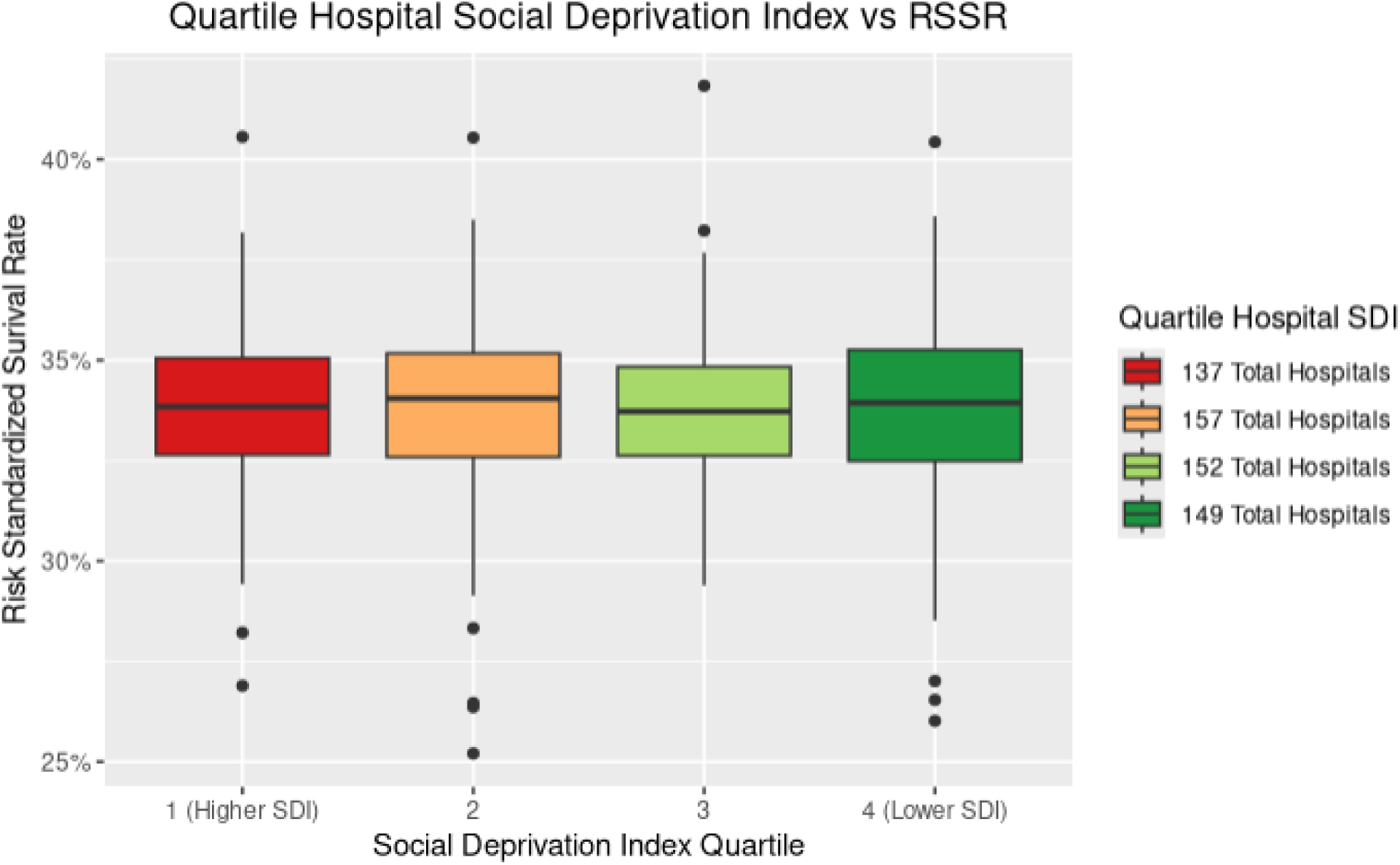
Risk-Standardized Post-resuscitation Survival by Quartile of Social Deprivation Index.

In the patient-level analysis where SDI quartile was included as an ordinal exposure variable, as compared to the highest SDI quartile (most deprivation), patients in lower SDI quartiles had higher rates of post-resuscitation survival (reference highest SDI (most deprivation) quartile, subsequent quartiles aOR 1.13, 95% CI 1.04-1.22, p<0.01, aOR 1.14, 95% CI 1.04-1.24, p<0.01, and lowest SDI (least deprivation) quartile aOR 1.13, 95% CI 1.03-1.23, p<0.01).

### Post-resuscitation Care Processes

A total of 98,679 (47.8%) patients were included in the analysis of the early fever process metric. In this subcohort, fever, including 24 hours of early death, occurred in 41,129 (41.7%) patients with rates across hospitals ranging from 9.5% to 85.7%. There was no monotonic association between hospital average rate of fever and RSSR quartile (p=0.43, Table 1). In the sensitivity analysis examining fever alone and without accounting for the competing risk of death within 24 hours, fever occurred in 21,308 (21.6%) patients with rates across hospitals ranging from 0% to 60.0%. As in the primary analysis, there was no monotonic association between hospital averaged rate of fever and RSSR quartile (p=0.65, Table 1).

**Table 1:** Rates of Early Fever and Early Death by Quartile of Risk-Standardized Post-Resuscitation Survival and Social Deprivation Index.

| Characteristic | Quartile 1 (Worst, lower RSSR, higher SDI) | 2 | 3 | 4 (Better, higher RSSR, lower SDI) | p-Value |
| --- | --- | --- | --- | --- | --- |
| **RSSR** |  |  |  |  |  |
| Fever | 43.7% | 43.5% | 44.3% | 44.9% | 0.43 |
| Fever (sensitivity analysis) | 21.6% | 20.9% | 21.3% | 20.8% | 0.65 |
| Early Death | 59.5% | 54.1% | 51.4% | 47.7% | <0.01 |
| **SDI** |  |  |  |  |  |
| Fever (early death) | 44.4% | 45.3% | 44.8% | 41.5% | 0.03 |
| Fever (sensitivity analysis) | 22.2% | 22.0% | 21.7% | 18.8% | 0.01 |
| Early Death | 51.2% | 52.7% | 54.0% | 54.4% | <0.01 |

Early death (≤72 hours) occurred in 94,048 (47.1%) patients, with rates of early death by hospital quartile similarly provided in Table 1. Hospitals in better RSSR quartiles had lower rates of early death (p<0.01).

Hospitals in higher SDI quartiles (most deprivation) had modestly higher rates of fever in both the primary and sensitivity analyses. Hospitals in higher SDI quartiles had higher rates of early death (Table 1).

## Discussion

The results of the present study demonstrate substantial hospital-to-hospital variation in RSSR for IHCA patients who survived the initial resuscitation. This variation exists upon a backdrop of stagnant or even worsening post-resuscitation survival over time, contrasting with improved overall IHCA survival over the past two decades. Numerous patient-level factors were associated with lower post-resuscitation survival at the patient level, including Black race, Hispanic ethnicity, and receiving care at a hospital with a higher social deprivation index. While the effect size of the association was modest, these findings highlight post-resuscitation care as a potential source of persistent disparity in cardiac arrest survival. In the exploratory process measure analysis, early death was more common in hospitals in worse RSSR quartiles and in hospitals in higher social deprivation quartiles. Early fever was more common in hospitals in higher social deprivation quartiles.

Post-resuscitation care has been recognized by the American Heart Association as a key link in the IHCA chain of survival.^22,23^ Nevertheless, we found no improvement (and possibly some worsening) in the adjusted odds of post-resuscitation survival over time in this study. While overall IHCA survival in the U.S. improved from 16.9% in 2000 to 26.7% in 2019,^2^ the primary driver appears to have been improvements in ROSC from over the same decades.^24^ Notably, we observed a substantially lower odds of post-resuscitation survival in 2021, likely related to the COVID-19 pandemic, and the odds of post-resuscitation survival have not returned to pre-pandemic levels in the following years (2022 to 2024). In a prior study including GWTG^®^-R data from years 2015-2018, Girotra et. al. found a similar overall rate of risk-adjusted post-resuscitation survival as in the present study. In that analysis, risk-adjusted post-resuscitation survival was tightly associated with overall hospital risk-standardized survival (including both initial IHCA survivors and non-survivors) but was not associated with acute resuscitation survival.^25^ This mirrors our findings where rates of acute resuscitation survival (ROSC) were similar across quartiles of risk-adjusted post-resuscitation survival.

We observed substantial variation in post-resuscitation RSSR, with many hospitals shifting in performance quartiles over time. The reason(s) for variation in post-resuscitation outcomes is not immediately clear, but the variation itself suggests a potential area for additional research and improvement. Potential contributors to variation in post-resuscitation care may include a less intensive focus on the post-resuscitation phase from quality improvement registries, therapeutic nihilism related to clinician expectations of outcome and potential self-fulfilling prophecy, and lack of clarity regarding optimal treatment approaches. Randomized controlled trials of post-arrest interventions, including blood pressure targets,^26,27^ oxygen targets,^28–30^ carbon dioxide targets,^28,31^ seizure prophylaxis,^32,33^ glucose control,^34^ prophylactic antibiotics,^35,36^ neuroprotective agents,^37–39^ corticosteroids,^40^ and recent trials of temperature control,^41–45^ have had mixed results which may contribute to variation in care practice. Advancing post-resuscitation care may require identifying and implementing the practices that distinguish higher-performing from lower-performing hospitals. The Hospital Enhancement of Resuscitation Outcomes for In-hospital Cardiac Arrest (HEROIC) studies successfully leveraged hospital-level variation to identify acute IHCA resuscitation practices associated with improved outcomes through qualitative interviews with staff at high and low-performing institutions.^46–48^ However, the HEROIC studies focused more on the resuscitation phase rather than post-arrest care; a similar approach for the post-ROSC period could help uncover institutional strategies that drive higher survival rates, generating actionable targets for improvement.

One potential contributor to hospital-to-hospital variation in post-resuscitation RSSR is the social deprivation index of the hospital area. We hypothesized that hospitals with more deprivation may lack the advanced expertise and equipment to implement often resource-intensive post-resuscitation therapies needed for temperature management, coronary catheterization, and neuroprognostication. In one study, Gonuguntla et al. found a > 2-fold difference in mortality between United States counties in the highest and lowest social vulnerability using the Social Vulnerability Index.^49^ Agerström et al. found that in a Swedish cohort low socioeconomic status was associated with delayed CPR and lower survival for IHCA.^50^ In the present study, quartile of SDI was a modest predictor of post-resuscitation survival at the patient level—although there was no association identified between quartile of hospital SDI and RSSR at the hospital level. Our findings highlight social deprivation as one of many potential drivers of variation in hospital post-resuscitation care performance, but also reflect that area-level deprivation explains only a small fraction of between-hospital differences; continued multilevel work incorporating case-mix, resources, and care processes is needed to clarify mechanisms and identify actionable targets.

Evaluating and tracking post-resuscitation process measures may help hospitals improve outcomes for patients who survive the initial resuscitation. In this study, we focused on two such measures, early fever and early death after initial resuscitation; selected both for their clinical relevance and availability within the GWTG^®^-R database. These measures are not only readily measurable and potentially modifiable, but they also reflect important aspects of post-arrest care. The AHA recommends maintaining a temperature between 32°C and 37.5°C after resuscitation, and early death prior to the opportunity for comprehensive prognostication may reflect guideline discordant care.^20,21,51^ In this study, we found that hospitals with higher proportions of early death (early death being used as a proxy for early withdrawal of life-sustaining therapy) were also in worse RSSR quartiles and higher social deprivation index quartiles. Hospitals in higher SDI quartiles (more deprivation) also had higher average rates of post-resuscitation early fever. While these findings should be considered exploratory only, they highlight how tracking post-resuscitation process metrics alongside intra-arrest metrics may give a fuller picture of performance and help identify modifiable opportunities for improvement.

Our study has limitations. First, we only included hospitals contributing data to the GWTG^®^-R registry, which limits generalizability. Second, a hospital’s SDI may not reflect the case-mix for institutions where a high volume of patients are transferred from outsize zip codes. Third, the large number of arrests in the GWTG^®^-R registry may allow for statistically significant associations that have a small effect size and are not clinically meaningful. Small effects should be interpreted with caution. Fourth, the process metrics analyzed are surrogates for guideline-concordant therapy and may not accurately reflect true adherence metrics. In addition, the fields required to assess for early fever are optional in the database, resulting in missingness.

## Conclusions

We found substantial hospital-to-hospital variation in RSSR against a backdrop of stagnant, or worsening, post-resuscitation outcomes over time despite overall IHCA survival gains. Black and Hispanic patients and those treated at hospitals serving more socially deprived areas had modestly lower post-resuscitation survival. Our findings highlight the post-resuscitation phase of care as an important area of research and improvement focus to save lives after IHCA.

## Data Availability

Data is available through the American Heart Association Get With The Guidelines-Resuscitation

